# Automated Multisource Electronic Frailty Index in Acute Ischemic Stroke: Development and Clinical Utility

**DOI:** 10.64898/2026.07.01.26357083

**Authors:** Lin Zhi Zheng Shawn, Foo Wei Ting, Ng Yee Sien, Chang Hui Meng, Laura Tay, Deidre Anne De Silva

## Abstract

**Background:** Frailty is common in acute ischemic stroke (AIS) and predicts poor outcomes, but is not routinely captured in acute stroke care. Manual frailty tools are difficult to apply consistently in busy inpatient settings, while existing electronic frailty indices (eFIs) often rely on limited data modalities. We developed a scalable pre-stroke electronic frailty index (eFI) using multisource electronic medical record (EMR) data and evaluated its clinical utility.

**Methods:** We conducted a retrospective cohort study of AIS admissions to Singapore General Hospital from July 1, 2024, to January 31, 2025. A fully automated pipeline derived an eFI from EMR data over a 3-year lookback period, incorporating ICD-10 codes, vital signs, anthropometry, laboratory results, medications, and free-text documentation processed using artificial intelligence–augmented extraction of predefined, clinically interpretable deficits. Candidate variables were screened using a validated 10-step frailty index framework and refined by multidisciplinary expert consensus.

**Results:** Among 501 AIS cases, the pipeline generated 75 candidate variables and a final 33-variable eFI, with scores derived for 492 cases (98.2%). Frail patients had greater premorbid disability, higher stroke severity, longer hospitalization, greater rehabilitation use, worse discharge disability, higher 30-day readmission, and higher cumulative post-discharge mortality. In multivariable analyses adjusted for age, sex, NIHSS, premorbid mRS, and reperfusion therapy, each 0.1-unit increase in eFI was associated with mortality beyond 90 days after discharge (adjusted HR, 1.47; 95% CI, 1.19–1.81), 30-day readmission (adjusted OR, 1.91; 95% CI, 1.43–2.59), longer hospital stay (β, 2.8 days; 95% CI, 1.4–4.2), and discharge to inpatient rehabilitation rather than home (adjusted RRR, 1.66; 95% CI, 1.27–2.16).

**Conclusions:** In a well-documented acute stroke service supported by comprehensive longitudinal EMR data, automated multisource eFI derivation was feasible and clinically informative in AIS, capturing baseline vulnerability beyond conventional stroke measures and supporting frailty-informed risk stratification and discharge planning.

## Introduction

Frailty, a state of reduced physiological reserve and vulnerability to stressors, is common in acute ischemic stroke (AIS), with reported prevalence estimates of approximately 25% to 67%^1,2^. Beyond its geriatric relevance, frailty has direct implications for stroke systems of care because of its associations with increased mortality, disability and length of admission^2–8^. Accurate and early identification of frail AIS patients is thus clinically important for informing prognostic discussions, anticipating rehabilitation and post-acute care needs, and guiding individualized discharge planning. Despite its clinical relevance, frailty remains difficult to assess reliably in acute stroke care. Conventional tools such as the Clinical Frailty Scale (CFS) are informative but require comprehensive assessment of premorbid function, often relying on patient or caregiver collateral that may not be immediately available or complete in time-sensitive acute stroke settings. In addition, aphasia, impaired consciousness, neglect, and motor deficits may further limit bedside assessment, while inter-rater variability and acute illness severity can affect the consistency and reliability of such tools^9,10^.

Electronic frailty indices (eFIs), derived from routinely collected electronic medical record (EMR) data using deficit accumulation principles, offer a scalable alternative. Several eFI models have been developed and validated in general populations and primary care settings, demonstrating utility for risk stratification and outcome prediction^11–17^. More recently, adaptations of eFI approaches have been explored in hospitalized and stroke populations. However, existing models are typically limited by labor-intensive data curation, reliance on administrative coding, restricted data modalities or complications arising during the index admission^18–22^. As a result, they may inadequately capture the multidimensional and dynamic nature of frailty preceding the index stroke, and their applicability as an early standardized measure within acute stroke workflows remains constrained.

To address these gaps, our study had two objectives. First, we aimed to develop an automated pre-stroke eFI integrating multi-source EMR structured data with free-text clinical documentation to provide a comprehensive representation of baseline frailty in AIS. Second, we aimed to examine the clinical utility of the derived eFI by evaluating its association with prespecified real-world stroke outcomes, including healthcare utilization, functional status, discharge disposition, readmission, and mortality. We hypothesized that our multi-source eFI would be feasible to derive at scale and would capture clinically meaningful vulnerability not fully reflected by conventional stroke measures such as age, stroke severity, or premorbid disability. By emphasizing automation, data completeness, and alignment with routine stroke workflows, our study sought to establish a scalable framework for frailty-informed risk stratification, prognostication, discharge planning, and future EMR-based clinical decision support systems.

## Methods

### Study Design and Population

We conducted a retrospective cohort study of consecutive patients admitted to the Singapore General Hospital Neurology Department between July 1, 2024, and January 31, 2025. Patients were identified from a clinically maintained acute stroke audit. Inclusion in the audit was based on a final hospital discharge diagnosis of acute ischemic stroke confirmed by the treating neurology specialist using clinical assessment and relevant neuroimaging findings.

### Ethics Approval and Data Governance

This retrospective study was reviewed and approved under the SGH institutional data governance and trusted third-party de-identification process for the use of anonymized data. The requirement for written informed consent was waived because the study used retrospectively collected anonymized data with no participant contact. Data de-identification was performed by the SGH Health Services Research Unit, acting as the institutional trusted third party, before release of the analysis dataset to the study team. All study procedures were conducted in accordance with institutional guidelines. One author (SLZZ) had full access to all anonymized data used in the study and takes responsibility for the integrity of the data and the accuracy of the data analysis.

### Data Sources

SGH is part of SingHealth, Singapore’s largest public healthcare group, which uses an integrated longitudinal EMR system across acute hospitals, community hospitals, national specialty centers and primary care clinics. EMR data recorded across SingHealth institutions during the 3-year lookback period preceding index stroke admission were extracted, including clinical diagnoses, vital signs and anthropometric measurements, laboratory investigations, medication records and free-text clinical documentation. Where applicable, diagnostic variables were derived from ICD-10 codes recorded across inpatient and outpatient encounters supplemented by the latest laboratory values, medication use, and specialist encounters prior to the AIS index admission to improve clinical specificity (**Table S1**).

### Data Processing and Feature Extraction

Structured data underwent unit standardization, range checks, outlier handling, and temporal alignment. Free-text clinical documentation was processed using a combination of rule-based algorithms and artificial intelligence (AI)-augmented feature extraction incorporating large language models to identify predefined frailty-relevant deficits. To minimize “black box” or unsupported classification, extraction was limited to clinically interpretable variables with explicit coding rules. Candidate variables were mapped to transparent operational definitions and reviewed by a multidisciplinary expert panel before inclusion in the final eFI (**Table S2**).

Feature extraction captured functional and geriatric syndromes (e.g., falls, incontinence), cognitive and behavioral indicators, healthcare utilization patterns, physiological abnormalities documented in clinical notes and derangements in laboratory biomarkers. Medication data were used to identify polypharmacy, high-risk medication classes, and treatment proxies for selected chronic diseases, such as bisphosphonate use for osteoporosis.

### eFI Construction

Candidate variables were evaluated using a validated 10-step frailty index framework^23^, including assessment of missingness, prevalence, redundancy, and age association, as well as evaluation of expected frailty index properties.

Variables were further refined through multidisciplinary expert consensus involving stroke neurologists, geriatricians, and rehabilitation specialists. While most variables demonstrated the expected positive association with age, selected clinically established deficits (diabetes, abnormal weight, malignancy, and chronic kidney disease) were retained despite lack of age correlation, consistent with frailty index principles that prioritize biological relevance over cohort-specific statistical associations.

The full derivation workflow, from multi-source EMR extraction to candidate deficit screening, expert consensus review, and final eFI construction, is summarized in **Fig 1**. The eFI was calculated as the proportion of deficits present out of the total assessed, generating a continuous score from 0 to 1, with higher values indicating greater frailty (**Fig S1**). Given the study focus on identifying patients with clinically significant vulnerability, individuals with eFI <0.25 (encompassing both robust and mild/pre-frail states)^12,14,24^ were analyzed as a single comparison group. This approach prioritizes discrimination of a clinically meaningful frail subgroup in the acute stroke setting.

**Figure 1.**
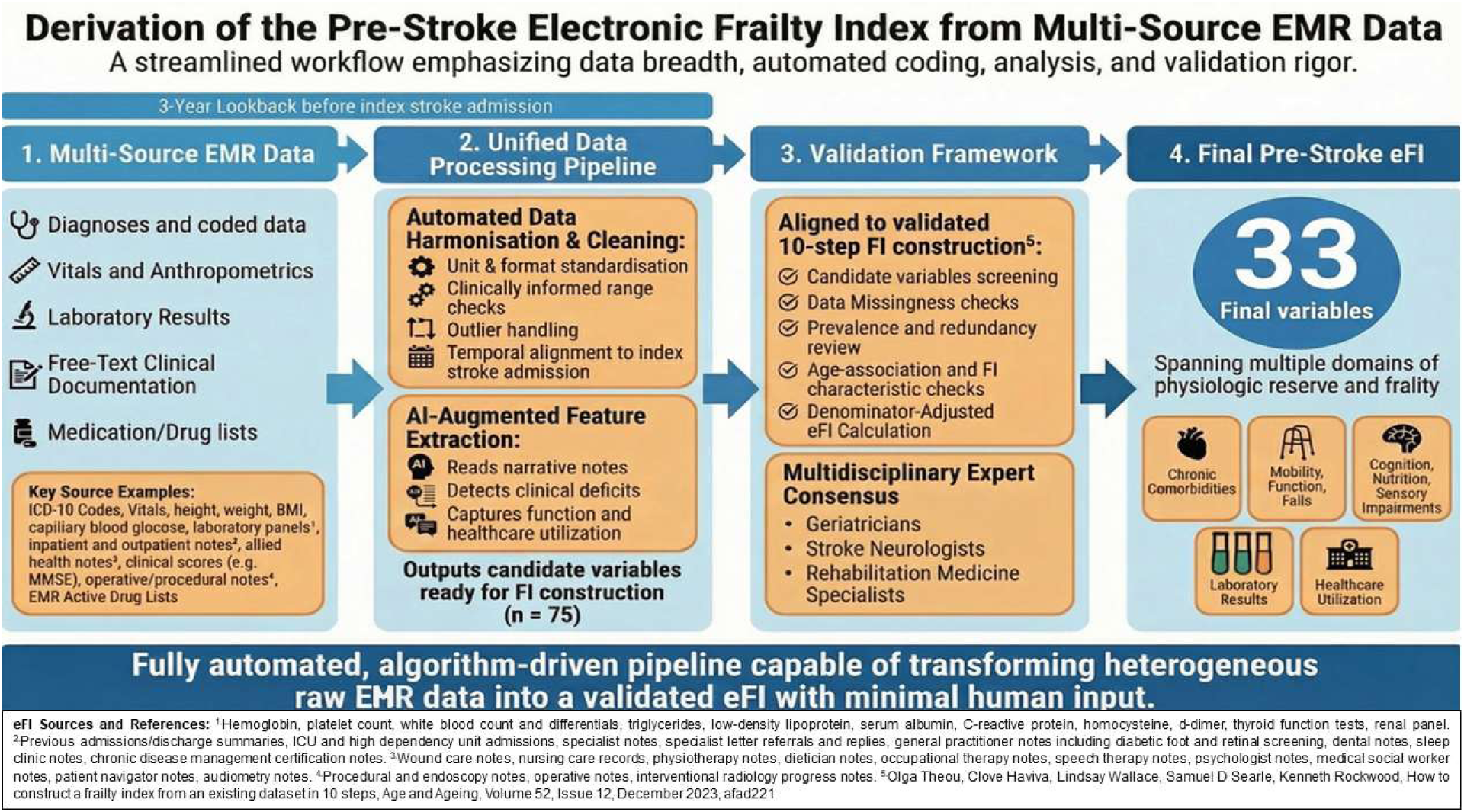
Derivation workflow for the stroke electronic frailty index (eFI). Abbreviations: AI, Artificial Intelligence; FI, Frailty Index. Multi-source EMR data from the 3 years preceding index stroke admission were integrated through an automated pipeline incorporating structured-data harmonization and AI-assisted free-text feature extraction. Candidate frailty deficits were screened using a validated 10-step frailty index construction framework and refined through multidisciplinary expert consensus, yielding a final 33-variable stroke eFI spanning multiple domains of physiologic reserve and frailty.

### Outcomes

Outcomes were prespecified to reflect key stages of the acute stroke pathway, including:

1. **Stroke presentation and classification** comprising clinical stroke severity assessed using the National Institutes of Health Stroke Scale (NIHSS) on admission, clinical stroke syndrome classified using the Oxfordshire Community Stroke Project (OCSP) criteria, and etiological subtype classified using the Trial of Org 10172 in Acute Stroke Treatment (TOAST) criteria;
2. **Hyperacute treatment**, including use of intravenous thrombolysis and endovascular thrombectomy (EVT);
3. **In-hospital course**, including length of stay and utilization of inpatient and post-acute stroke rehabilitation services;
4. **Early post-stroke outcomes**, including recurrent stroke within 90 days and discharge disposition (home, inpatient rehabilitation, or other facility);
5. **Functional outcomes**, including modified Rankin Scale (mRS) at discharge and requirement for assistance with activities of daily living (ADLs) including tube feeding, dressing, toileting, and ambulation;
6. **Healthcare utilization and survival**, including 30-day readmission and all-cause mortality during follow-up.

Subgroup analyses were performed among patients treated with reperfusion therapy. In the EVT subgroup, procedural outcomes included procedure duration, final modified Treatment in Cerebral Infarction (mTICI) reperfusion grade, and NIHSS at 24–48 hours after EVT. Safety outcomes included any hemorrhagic transformation on follow-up imaging, parenchymal hematoma type 1 or 2 according to the European Cooperative Acute Stroke Study III (ECASS III) classification, and documented procedural complications. Intravenous thrombolysis subgroup analyses were descriptive because of the small number of treated frail patients and limited complete safety outcome data. Outcomes included door-to-needle time as a treatment-process measure and recorded treatment-related complications as safety measures.

### Outcome Ascertainment and Follow-up

Clinical outcomes were ascertained retrospectively from the integrated SingHealth EMR through the administrative data cut-off of April 15, 2026. Thirty-day readmission was assessed during the 30 days after discharge date from the index AIS admission, while recurrent stroke was assessed during the 90 days after the index admission date. All included patients had at least 90 days of potential observation before the administrative data cut-off.

All-cause mortality status was obtained through deterministic person-level linkage between the SingHealth electronic medical record and national death-registration records using a unique national identifier. Linkage was performed centrally by the institutional data custodian before de-identification and release of the anonymized analysis dataset to the study team. The investigators did not perform the linkage or independently evaluate linkage quality. Follow-up for mortality commenced on the date of discharge and continued until death or administrative censoring on April 15, 2026. Mortality status was available for all eligible patients through the administrative data cut-off; therefore, there was no loss to follow-up for mortality ascertainment. No direct participant contact was undertaken.

### Statistical Analysis

Continuous variables were summarized as median (interquartile range), and categorical variables as counts (percentages). Between-group comparisons used the Wilcoxon rank-sum test or Student t test for continuous variables, as appropriate, and the χ² test or Fisher exact test for categorical variables. For descriptive comparisons, NIHSS was categorized as minor stroke (0–5), moderate stroke (6–15), moderate-to-severe stroke (16–20), and severe stroke (21–42); NIHSS was modeled as a continuous variable in multivariable analyses.

To preserve statistical power and reduce reliance on a single frailty threshold, eFI was modeled continuously per 0.1-unit increase. Models were adjusted for age, sex, NIHSS on presentation, premorbid mRS, EVT, and intravenous thrombolysis. Complete-case analysis was used for the variables included in each model, and no imputation was performed. Denominators differing from the total sample size reflect outcome-specific or covariate-specific missingness.

Associations between eFI and binary clinical outcomes were assessed using multivariable logistic regression. Acute hospital length of stay was evaluated using multivariable linear regression. Discharge destination was evaluated using multinomial logistic regression, with discharge home as the reference category.

Associations with mortality were evaluated using Cox proportional hazards regression. The proportional hazards assumption was assessed using scaled Schoenfeld residuals for each covariate and for the model globally; P<0.05 was considered evidence of nonproportional hazards. Because nonproportional hazards were identified for eFI, age, and premorbid mRS, a piecewise Cox model was fitted to accommodate their time-varying associations with mortality. The 30- and 90-day boundaries were selected after examination of the proportional hazards diagnostics as clinically interpretable post-discharge periods, resulting in intervals of 0–30 days, >30–90 days, and >90 days after discharge. Interval-specific effects were estimated for eFI, age, and premorbid mRS, whereas the effects of sex, NIHSS, EVT, and intravenous thrombolysis were constrained to remain constant across intervals. Robust standard errors clustered by patient identifier were used to account for multiple interval-specific records contributed by the same patient.

Linearity of continuous predictors in the logit was evaluated for logistic regression models, and linear regression assumptions were assessed through examination of model residuals. Multicollinearity was assessed using generalized variance inflation factors (GVIF); all adjusted GVIF values were below 2, indicating no evidence of meaningful multicollinearity. EVT subgroup analyses were considered exploratory because of the limited sample size and number of outcome events. All tests were 2-sided, with P<0.05 considered statistically significant. Analyses were conducted using R version 4.4.0.

### Data Availability

Individual-level data are not publicly available because of institutional data-governance and patient-confidentiality requirements. Detailed variable definitions and coding algorithms are provided in the Supplemental Material. Analytical code may be made available upon reasonable request to the corresponding author, subject to institutional approval.

## Results

### Feasibility and eFI Characteristics

A total of 501 AIS cases were included, with eFI successfully derived for 492 (**98.2%**). The eFI could not be derived for 9 patients because the available EMR data were insufficient to calculate the index. The final eFI comprised 33 variables (**Table 1**) derived from an initial 75 candidate variables (**Table S2**). The eFI distribution was right-skewed and increased with age, consistent with expected frailty index properties, with generally higher values observed in females^23^ (**Fig S2 and S3**).

**Table 1.** Clinical Domains and Representative Variables of the Stroke eFI.

| Clinical Domain | Representative variables |
| --- | --- |
| Cardiometabolic | Heart failure/valvular heart disease/cardiac arrhythmias, ischemic heart disease, diabetes, thyroid and other endocrine disorders, abnormal BMI, malnutrition, CKD/dialysis |
| Gastrointestinal | GI and HPB disease, dysphagia and abdominal symptoms |
| Sensory Impairment, mobility and social vulnerability | Hearing impairment, visual impairment, history of falls, poor social support requiring MSW intervention |
| Skin and continence | Pressure ulcers, urinary and/or bowel incontinence |
| High risk medication burden | Benzodiazepine use |
| Geriatric complexity | Complex geriatric syndromes requiring specialist geriatrician management |
| Hematologic and Oncologic Conditions | Anaemia, history of cancer |
| Neurocognitive and Neurological Disorders | Dementia, altered mental status, giddiness, history of neurosurgical conditions or procedures |
| Respiratory | Chronic lung disease, dyspnoea, asthma |
| Musculoskeletal/Vascular | Orthopaedic conditions and surgeries, chronic pain disorders, osteoporosis and bisphosphonate use, peripheral vascular disease |
| Rehabilitation need and Multimorbidity | Prior multidisciplinary rehabilitation requirement, history of multiple chronic diseases requiring medical follow up |

### Baseline Cohort Characteristics

Among patients with derived eFI scores, 182 of 492 (37.0%) were classified as frail (eFI ≥0.25). Frail patients were older than non-frail patients (age, 77 [72–83] versus 74 [69–81] years; P<0.001) and were more likely to have poor premorbid functional status (mRS ≥3, 37% versus 15%; p<0.001). They also had a higher burden of vascular comorbidities, including atrial fibrillation (34% versus 23%; p=0.014), previous stroke (34.6% versus 24.3%; p=0.024), hypertension (86% versus 73%; p=0.002), hyperlipidemia (77% versus 62%; p<0.001), and diabetes (52% versus 35%; p<0.001). Sex distribution, body mass index (BMI), alcohol use, smoking history, and ethnicity were similar between groups (**Table 2**).

**Table 2.**
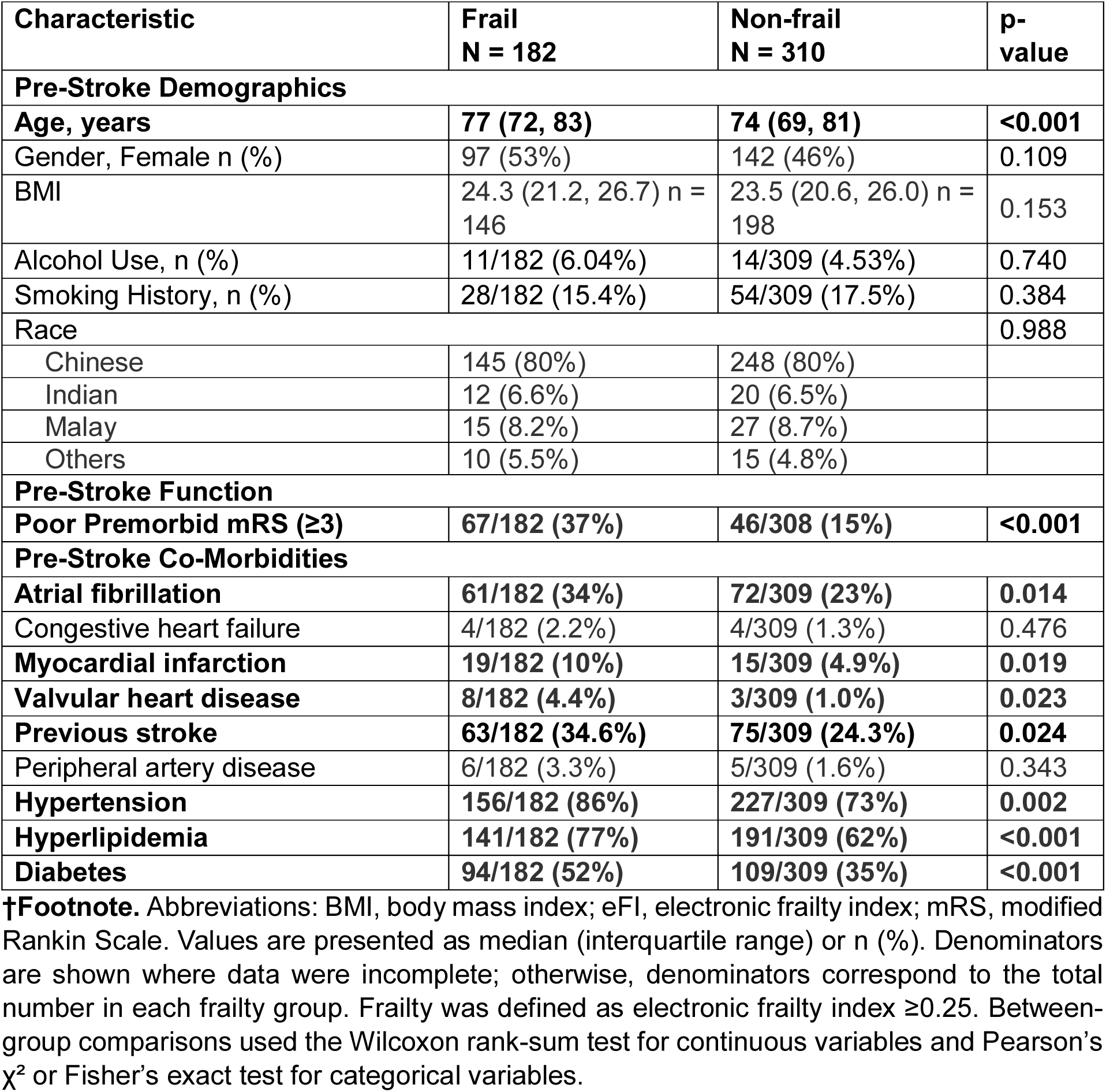
Baseline Characteristics of participants stratified by Frailty Status.

| Characteristic | Frail<br>N = 182 | Non-frail<br>N = 310 | p-value |
| --- | --- | --- | --- |
| **Pre-Stroke Demographics** |  |  |  |
| **Age, years** | **77 (72, 83)** | **74 (69, 81)** | **<0.001** |
| Gender, Female n (%) | 97 (53%) | 142 (46%) | 0.109 |
| BMI | 24.3 (21.2, 26.7) n = 146 | 23.5 (20.6, 26.0) n = 198 | 0.153 |
| Alcohol Use, n (%) | 11/182 (6.04%) | 14/309 (4.53%) | 0.740 |
| Smoking History, n (%) | 28/182 (15.4%) | 54/309 (17.5%) | 0.384 |
| Race |  |  | 0.988 |
| Chinese | 145 (80%) | 248 (80%) |  |
| Indian | 12 (6.6%) | 20 (6.5%) |  |
| Malay | 15 (8.2%) | 27 (8.7%) |  |
| Others | 10 (5.5%) | 15 (4.8%) |  |
| **Pre-Stroke Function** |  |  |  |
| **Poor Premorbid mRS (≥3)** | **67/182 (37%)** | **46/308 (15%)** | **<0.001** |
| **Pre-Stroke Co-Morbidities** |  |  |  |
| **Atrial fibrillation** | **61/182 (34%)** | **72/309 (23%)** | **0.014** |
| Congestive heart failure | 4/182 (2.2%) | 4/309 (1.3%) | 0.476 |
| **Myocardial infarction** | **19/182 (10%)** | **15/309 (4.9%)** | **0.019** |
| **Valvular heart disease** | **8/182 (4.4%)** | **3/309 (1.0%)** | **0.023** |
| **Previous stroke** | **63/182 (34.6%)** | **75/309 (24.3%)** | **0.024** |
| Peripheral artery disease | 6/182 (3.3%) | 5/309 (1.6%) | 0.343 |
| **Hypertension** | **156/182 (86%)** | **227/309 (73%)** | **0.002** |
| **Hyperlipidemia** | **141/182 (77%)** | **191/309 (62%)** | **<0.001** |
| **Diabetes** | **94/182 (52%)** | **109/309 (35%)** | **<0.001** |

†Footnote. Abbreviations: BMI, body mass index; eFI, electronic frailty index; mRS, modified Rankin Scale. Values are presented as median (interquartile range) or n (%). Denominators are shown where data were incomplete; otherwise, denominators correspond to the total number in each frailty group. Frailty was defined as electronic frailty index ≥0.25. Between-group comparisons used the Wilcoxon rank-sum test for continuous variables and Pearson's $\chi^2$ or Fisher's exact test for categorical variables.

### Index Stroke Characteristics

Frail patients presented with more severe strokes, with a higher proportion classified as severe by NIHSS compared with non-frail patients (22% versus 13%; overall p=0.014). They were less likely to receive intravenous thrombolysis (1.6% versus 7.1%; p=0.008), while EVT use was similar between groups (20% versus 20%; p=0.930). There were no significant differences in clinical stroke syndrome by OCSP classification or etiological subtype by TOAST classification. Frail patients were more likely to require dedicated stroke rehabilitation during the acute admission (99% versus 95%; p=0.037) and post-acute care phase (64% versus 48%; p=0.003) (**Table 3**).

**Table 3.** Index Acute Ischemic Stroke Admission Characteristics of participants stratified by Frailty Status.

| Characteristic | Frail<br>N = 182 | Non-frail<br>N = 310 | p-value |
| --- | --- | --- | --- |
| **Severity of Stroke based on NIHSS** |  |  | **0.014** |
| Minor stroke (NIHSS 0-5) | 87/172 (51%) | 191/295 (65%) |  |
| Moderate stroke (NIHSS 6-15) | 37/172 (22%) | 54/295 (18%) |  |
| Moderate to Severe stroke (NIHSS 16-20) | 11/172 (6.4%) | 13/295 (4.4%) |  |
| Severe Stroke (NIHSS 21-42) | 37/172 (22%) | 37/295 (13%) |  |
| OCSP classification |  |  | 0.426 |
| LACI | 68 (37%) | 139 (45%) |  |
| PACI | 64 (35%) | 102 (33%) |  |
| POCI | 25 (14%) | 38 (12%) |  |
| TACI | 25 (14%) | 31 (10%) |  |
| TOAST classification |  |  | 0.155 |
| Cardioembolism | 55 (30%) | 64 (21%) |  |
| Large-artery atherosclerosis | 27 (15%) | 54 (17%) |  |
| Small-vessel occlusion | 30 (16%) | 62 (20%) |  |
| Undetermined etiology | 70 (38%) | 130 (42%) |  |
| **Intravenous Thrombolysis given, n (%)** | **3 (1.6%)** | **22 (7.1%)** | **0.008** |
| EVT done, n (%) | 39 (20%) | 62 (20%) | 0.930 |
| **Acute inpatient care included dedicated stroke rehabilitation** | **124/125 (99%)** | **209/221 (95%)** | **0.037** |
| **Post acute care included dedicated stroke rehabilitation** | **80/125 (64%)** | **105/221 (48%)** | **0.003** |

‡Footnote. Abbreviations: NIHSS, National Institutes of Health Stroke Scale; OCSP, Oxfordshire Community Stroke Project; LACI, lacunar infarct; PACI, partial anterior circulation infarct; POCI, posterior circulation infarct; TACI, total anterior circulation infarct; TOAST, Trial of Org 10172 in Acute Stroke Treatment; EVT, endovascular thrombectomy. Values are presented as median (interquartile range) or n (%). Denominators are shown where data were incomplete; otherwise, denominators correspond to the total number in each frailty group. Frailty was defined as electronic frailty index $\geq 0.25$ . Between-group comparisons used the Wilcoxon rank-sum test for continuous variables and Pearson's $\chi^2$ , Fisher's exact test, or Fisher's Exact Test for Count Data with simulated p-value (based on 10000 replicates).

### Hospitalization and Early Outcomes

Frail patients had longer acute hospital stays than non-frail patients (10 [6–22] versus 6 [3–11] days; P<0.001) and worse discharge disability (mRS, 4 [3–4] versus 3 [1–4]; P<0.001). They were less often discharged home (53% versus 68%) and more frequently transferred to inpatient rehabilitation (44% versus 25%; overall p=0.004). Frail patients also had higher 30-day readmission (22% versus 5.9%; p<0.001) and a higher proportion of all-cause post-discharge deaths (33% versus 19%; p<0.001). There were only 10 cases of 90-day recurrent stroke in the overall cohort, with no significant difference between frail and non-frail groups. (**Table 4**).

**Table 4.** Stroke Outcomes Stratified by Frailty Status.

| **Stroke Outcome** | **Frail**<br>**N = 182** | **Non-frail**<br>**N = 310** | **p-**<br>**value** |
| --- | --- | --- | --- |
| **mRS score on discharge** | **4.00 (3.00, 4.00) n = 181** | **3.00 (1.00, 4.00) n = 305** | **<0.001** |
| Persistent dysphagia requiring tube feeding at discharge | 23 (19%) | 31 (14%) | 0.261 |
| **Required assistance with dressing at discharge** | **27 (39%)** | **23 (18%)** | **0.002** |
| **Required assistance with toileting at discharge** | **27 (39%)** | **23 (18%)** | **0.002** |
| New stroke within 90 days of admission | 6/125 (4.8%) | 4/221 (1.8%) | 0.178 |
| **Acute hospital length of stay, days** | **10 (6, 22)** | **6 (3, 11)** | **<0.001** |
| **30-day hospital readmission** | **40/180 (22%)** | **18/305 (5.9%)** | **<0.001** |
| **All-cause post-discharge mortality** | **60 (33%)** | **58 (19%)** | **<0.001** |
| **Discharge destination** |  |  | **0.004** |
| Another acute care hospital | 0/125 (0%) | 2/221 (0.9%) |  |
| **Inpatient rehabilitation facility** | **55/125 (44%)** | **55/221 (25%)** |  |
| **Home** | **66/125 (53%)** | **150/221 (68%)** |  |
| Others | 0/125 (0%) | 2/221 (0.9%) |  |
| Inpatient Mortality | 4/125 (3.2%) | 10/221 (4.5%) |  |
| Residential facility (e.g. Nursing Home) | 0/125 (0%) | 2/221 (0.9%) |  |

**§Footnote.** Abbreviations: mRS, modified Rankin Scale. Values are presented as median (interquartile range) or n (%). Denominators are shown where data were incomplete; otherwise, denominators correspond to the total number in each frailty group. Frailty was defined as electronic frailty index $\geq 0.25$ . Between-group comparisons used the Wilcoxon rank-sum test for continuous variables and Pearson's $\chi^2$ or Fisher's exact test for categorical variables, as appropriate. Mortality percentages represent crude proportions of patients who died during available follow-up through the administrative data cut-off and are not fixed-time mortality risks. Time-to-event associations are presented in Table 5 and Table S3.

### Multivariable Association Between eFI and Key Stroke Outcomes

In multivariable analyses (**Table 5**), higher eFI was independently associated with several clinically important outcomes. Each 0.1-unit increase in eFI was associated with higher odds of 30-day readmission (adjusted OR, 1.91; 95% CI, 1.43–2.59; P<0.001), a greater likelihood of discharge to inpatient rehabilitation rather than home (adjusted RRR, 1.66; 95% CI, 1.27–2.16; P<0.001), and an adjusted mean increase of 2.8 days in acute hospital length of stay (95% CI, 1.4–4.2; P<0.001). In contrast, eFI was not independently associated with functional dependence at discharge (mRS ≥3; adjusted OR, 1.20; 95% CI, 0.92–1.56; P=0.183). Discharge mRS ≥3 was more strongly associated with NIHSS on presentation (adjusted OR per 1-point increase, 1.14; 95% CI, 1.08–1.21; P<0.001) and premorbid mRS (adjusted OR per 1-point increase, 2.67; 95% CI, 1.95–3.83; P<0.001).

**Table 5.** Multivariable Association Between eFI and Key Stroke Outcomes.

| Stroke Outcomes | Predictor | Estimate Type | Effect Estimate | 95% CI | p-value |
| --- | --- | --- | --- | --- | --- |
| **All-cause post-discharge mortality Beyond 90 days** | **eFI (per 0.1)** | **Adjusted HR** | **1.47** | **1.19, 1.81** | **<0.001** |
|  | **NIHSS (per 1)** |  | **1.09** | **1.06, 1.11** | **<0.001** |
|  | Premorbid mRS (per 1) |  | 1.19 | 0.98, 1.44 | 0.072 |
| **Discharge mRS $\geq 3$** | **eFI (per 0.1)** | **Adjusted OR** | 1.20 | 0.92, 1.56 | 0.183 |
|  | **NIHSS (per 1)** |  | **1.14** | **1.08, 1.21** | **<0.001** |
|  | **Premorbid mRS (per 1)** |  | **2.67** | **1.95, 3.83** | **<0.001** |
| **30-day readmission** | **eFI (per 0.1)** | **Adjusted OR** | **1.91** | **1.43, 2.59** | **<0.001** |
|  | NIHSS (per 1) |  | 0.98 | 0.94, 1.02 | 0.369 |
|  | **Premorbid mRS (per 1)** |  | **1.52** | **1.20, 1.92** | **<0.001** |
| **Acute hospital length of stay, days** | **eFI (per 0.1)** | **$\beta$ coefficient** | **2.8** | **1.4, 4.2** | **<0.001** |
|  | **NIHSS (per 1)** |  | **0.41** | **0.21, 0.61** | **<0.001** |
|  | Premorbid mRS (per 1) |  | -1.2 | -2.4, 0.05 | 0.061 |
| **Discharge to inpatient rehabilitation facility versus home** | **eFI (per 0.1)** | **Adjusted RRR** | **1.66** | **1.27, 2.16** | **<0.001** |
|  | NIHSS (per 1) |  | 1.04 | 1.00, 1.08 | 0.068 |
|  | Premorbid mRS (per 1) |  | 0.81 | 0.65, 1.01 | 0.065 |

**Footnote.** Abbreviations: HR, hazard ratio; OR, odds ratio; CI, confidence interval; RRR, relative risk ratio. For length of stay, estimates are $\beta$ coefficients in days. For discharge to inpatient rehabilitation versus home, estimates are adjusted relative risk ratios from multinomial logistic regression. All models were adjusted for age, sex, NIHSS on presentation, premorbid mRS, and reperfusion therapy use.

The complete-case dataset for the piecewise Cox model included 443 patients and 110 deaths. Eight of the 118 deaths recorded in the overall cohort were excluded from the adjusted survival analysis because of missing data for one or more model covariates. Among the deaths included in the model, 31 occurred during days 0–30, 15 during days 31–90, and 64 beyond 90 days after discharge. The association between eFI and post-discharge mortality varied over time. eFI was not significantly associated with mortality during the 0–30-day interval (adjusted HR per 0.1-unit increase, 0.73; 95% CI, 0.52–1.01; P=0.055) or the 31–90-day interval (adjusted HR, 1.04; 95% CI, 0.68–1.58; P=0.857). Beyond 90 days, each 0.1-unit increase in eFI was associated with a 47% higher hazard of death (adjusted HR, 1.47; 95% CI, 1.19–1.81; P<0.001). Among the other model covariates, NIHSS remained independently associated with mortality across follow-up (adjusted HR per 1-point increase, 1.09; 95% CI, 1.06–1.11; P<0.001). Age was associated with mortality only beyond 90 days, whereas premorbid mRS was not significantly associated with mortality in any follow-up interval. The full piecewise Cox model, including interval-specific estimates for age and premorbid mRS and constant effects for sex, NIHSS, EVT, and intravenous thrombolysis, is presented in **Table S3**.

### EVT Sub-Cohort Outcomes

Among patients undergoing EVT, frail patients had longer procedure duration than non-frail patients (28 [24–44] versus 24 [18–33] minutes; P=0.048), but final mTICI grade, 24–48-hour NIHSS, hemorrhagic transformation, and procedural complication rates were not significantly different between groups (**Table 6**). In adjusted exploratory analyses, eFI was not significantly associated with procedural complications but was inversely associated with ECASS III PH1/PH2 hemorrhage (**Table 7**).

**Table 6.** EVT Sub-Cohort Treatment Outcomes stratified by Frailty Status.

| Outcome | frail<br>N = 37 | non-frail<br>N = 62 | p-value |
| --- | --- | --- | --- |
| EVT duration (mins) | 28 (24, 44) n = 37 | 24 (18, 33) n = 50 | 0.048 |
| Post EVT mTICI score |  |  | 0.126 |
| 0 | 1/37 (2.7%) | 6/58 (10%) |  |
| 1 | 0/37 (0%) | 3/58 (5.2%) |  |
| 2a | 2/37 (5.4%) | 1/58 (1.7%) |  |
| 2b | 7/37 (19%) | 3/58 (5.2%) |  |
| 2c | 9/37 (24%) | 16/58 (28%) |  |
| 3 | 18/37 (49%) | 29/58 (50%) |  |
| NIHSS score at 24-48hrs post EVT | 7 (3, 14) n = 25 | 6 (3, 10) n = 35 | 0.588 |
| Imaging outcomes: Any haemorrhagic transformation | 18 (49%) | 30 (48%) | 0.980 |
| Any procedural complications | 2 (5.4%) | 11 (18%) | 0.123 |

**#Footnote.** Abbreviations: EVT, endovascular thrombectomy; mTICI, modified Treatment in Cerebral Infarction; NIHSS, National Institutes of Health Stroke Scale. Values are presented as median (interquartile range) or n (%). Denominators are shown where data were incomplete; otherwise, denominators correspond to the total number in each frailty group. Frailty was defined as electronic frailty index $\geq 0.25$ . Between-group comparisons used the Wilcoxon rank-sum test for continuous variables and Pearson's $\chi^2$ or Fisher's exact test for categorical variables, as appropriate. Analyses were restricted to patients who underwent EVT; results should be interpreted cautiously given incomplete outcome data and limited event numbers.

**Table 7.** Multivariable Association Between eFI and EVT Outcomes/Safety.

| **Outcome** | **Adjusted OR** | **95% CI** | **p-value** |
| --- | --- | --- | --- |
| **Procedural complications (No Obs. = 94)** |  |  |  |
| eFI (per 0.1) | 0.51 | 0.23, 1.00 | 0.066 |
| NIHSS (per 1) | 0.97 | 0.89, 1.05 | 0.474 |
| Premorbid mRS (per 1) | 0.77 | 0.34, 1.40 | 0.450 |
| **ECASS III PH1/PH2 (No Obs. = 44)** |  |  |  |
| **eFI (per 0.1)** | **0.22** | **0.05, 0.60** | **0.010** |
| **NIHSS (per 1)** | **1.14** | **1.03, 1.32** | **0.030** |
| Premorbid mRS (per 1) | 1.56 | 0.76, 3.30 | 0.216 |

**\*\*Footnote.** Abbreviations: OR, Odds Ratio; CI, Confidence Interval; ECASS III, European Cooperative Acute Stroke Study III; PH, parenchymal hematoma. Effect estimates are adjusted odds ratios from multivariable logistic regression models. All models were adjusted for age, sex, NIHSS on presentation, premorbid mRS, and intravenous thrombolysis use. Analyses were restricted to patients who underwent EVT with complete outcome and covariate data. Given the small subgroup and limited event numbers, estimates should be interpreted as exploratory.

### Intravenous Thrombolysis Sub-Cohort Outcomes

Further adjusted analysis of the intravenous thrombolysis subgroup was not performed because few frail patients received thrombolysis and complete safety outcome data were limited. Descriptively, door-to-needle time and recorded treatment-related safety events did not differ significantly by frailty status (**Table S4**).

## Discussion

In this real-world cohort of patients hospitalized with AIS, we developed an automated, multi-source EMR-derived pre-stroke eFI and demonstrated high feasibility, with scores generated for 98% of patients. Frailty was common, affecting 36% of the overall cohort. Although this prevalence is higher than the pooled estimate of 24.6% reported in a 2022 acute stroke meta-analysis^2^, it remains consistent with the substantial heterogeneity across stroke cohorts, frailty instruments, and study populations, as well as more recent older-AIS series reporting high frailty burden^25^. Consistent with prior systematic reviews^2,5^, observational studies^3,4,6,25,26^, and the 2025 World Stroke Organization scientific statement^27^, higher frailty in our cohort was associated with more severe stroke presentation and, in adjusted analyses, with higher longer-term mortality, readmission, longer hospitalization, and greater post-acute care needs. These findings support frailty as a clinically meaningful dimension of stroke vulnerability beyond chronological age.

The principal contribution of this study is a clinically scalable approach to ascertain pre-stroke frailty from routinely available longitudinal EMR data. Previous stroke studies have relied mainly on bedside frailty instruments, research-assembled indices, or administrative scores. Bedside tools may be difficult to apply consistently in time-sensitive stroke care^9,10^, whereas administrative measures may be limited by coding practices, restricted data modalities, or inclusion of index-admission complications^18–20,22^. Our eFI instead used a defined 3-year pre-stroke period and combined structured data with AI-augmented extraction of predefined, clinically interpretable deficits from free-text documentation. Constructed using deficit-accumulation principles, a validated 10-step framework^23^, and multidisciplinary expert consensus, it captured multiple domains relevant to frailty^28^ including comorbidity, function, cognition, nutrition, medication burden, social vulnerability, and healthcare utilization.

Frailty, acute stroke severity, and premorbid disability capture different dimensions of vulnerability. NIHSS reflects the neurological severity of the index stroke, whereas premorbid mRS reflects pre-existing functional dependence; neither fully captures physiological reserve^29,30^. In our cohort, discharge dependence was more strongly associated with NIHSS and premorbid mRS, whereas eFI remained associated with readmission, hospitalization burden, rehabilitation needs, and mortality beyond 90 days. The time-varying mortality association further supports this distinction: NIHSS remained associated with mortality throughout follow-up, whereas eFI was associated with mortality only beyond 90 days. This may reflect the greater influence of acute neurological injury on early outcomes^31,32^, with accumulated physiological vulnerability becoming more relevant during longer-term survival^33^. These findings suggest that eFI provides complementary prognostic information rather than duplicating conventional measures of stroke severity or premorbid disability.

Automated frailty ascertainment may complement clinical assessment when bedside evaluation is limited by aphasia, neglect, impaired consciousness, motor deficits, or unavailable collateral history. It could support risk stratification, prognostic discussions, rehabilitation planning, caregiver counselling, and post-acute care coordination. Frailty should not, however, be used alone to withhold evidence-based stroke therapies. In our exploratory reperfusion analyses, most EVT procedural and safety outcomes did not differ clearly by frailty status, consistent with evidence that frailty is associated with mortality among reperfusion-treated patients but not clearly with symptomatic intracranial hemorrhage or poststroke disability^34^. Conversely, the inverse association between eFI and ECASS III PH1/PH2 hemorrhage should not be interpreted as protective, as it may reflect small event numbers, lower thrombolysis use among frail patients, and treatment-selection effects. Larger multicenter studies are needed to clarify whether frailty modifies reperfusion benefit or risk, and how frailty should be incorporated into hyperacute stroke decision-making.

This study has several strengths. It used an automated multisource EMR pipeline drawing on longitudinal data captured across an integrated public healthcare cluster, rather than relying solely on index-admission data or administrative coding. The pipeline integrated structured and unstructured clinical data, achieved a high eFI derivation rate, applied a validated frailty index construction framework, and evaluated clinically relevant outcomes across the acute stroke pathway. Several limitations should also be acknowledged. First, although the EMR data spanned multiple healthcare settings within an integrated cluster, this was a single-center retrospective cohort, limiting generalizability, precluding causal inference, and leaving the possibility of residual or unmeasured confounding. Reliance on routinely collected data may also have led to misclassification of individual eFI deficits because of incomplete documentation or variable healthcare contact, despite predefined operational definitions and multidisciplinary expert review. Feasibility and performance may therefore differ across institutions with different documentation practices, data systems, and healthcare utilization patterns. Second, the piecewise Cox intervals were selected after review of proportional hazards diagnostics and should therefore be considered exploratory. Early interval estimates were also limited by relatively few deaths while complete-case mortality analysis may have introduced selection bias. Reperfusion analyses were similarly limited by small sample sizes and event numbers. Third, the eFI was not directly compared with contemporaneous clinician-assessed frailty measures, precluding formal criterion validation. Nonetheless, the expected distribution of eFI scores, association with age, and independent associations with mortality, readmission, length of stay, and discharge disposition support its construct and prognostic validity. External validation across healthcare systems is needed before routine clinical deployment.

## Conclusion

Using longitudinal multisource EMR data incorporating structured and free-text clinical documentation across an integrated healthcare cluster, automated pre-stroke eFI derivation was feasible in patients with AIS. The resulting eFI captured baseline vulnerability not fully reflected by age, stroke severity, or premorbid disability, and was independently associated with longer-term mortality, readmission, acute length of stay, and discharge to inpatient rehabilitation rather than home. These findings support further validation of EMR-derived frailty assessment as a clinically useful approach to identify vulnerable stroke patients early, inform prognostic discussions, anticipate rehabilitation and post-acute care needs, and guide individualized discharge planning.

## Clinical Perspective

### What Is New?

- A fully automated electronic frailty index was derived from multisource EMR data in 98% of patients hospitalized with acute ischemic stroke.
- The index integrates structured clinical data with clinically constrained extraction of predefined frailty deficits from free-text documentation over a 3-year lookback period from index stroke admission.
- Higher eFI was associated with longer-term mortality, readmission, length of stay, and discharge to inpatient rehabilitation after adjustment for age, stroke severity, premorbid disability, and reperfusion therapy.

### What Are the Clinical Implications?

- Automated frailty ascertainment may help clinicians identify vulnerable acute ischemic stroke patients early, particularly when aphasia, impaired consciousness, neglect, or motor deficits limit bedside frailty assessment.
- The eFI may support earlier rehabilitation referral, geriatric or allied health input, caregiver counselling, discharge planning, and post discharge follow-up for patients at higher risk of complex recovery.
- Because EMR-derived frailty assessment depends on local data availability and documentation practices, external validation is needed before routine deployment across institutions and EMR systems.

## Acknowledgments

The authors thank Mr Xu Yang, Senior Analyst at the Singapore General Hospital Health Services Research Unit, for support with trusted third-party data de-identification and preparation of the anonymized dataset. We also acknowledge the Singapore General Hospital Research Office, the Department of Neurology at the National Neuroscience Institute, Singapore General Hospital Campus, and the clinical and administrative teams involved in maintaining the acute stroke audit data that supported this study.

## Use of Artificial Intelligence

During preparation of this manuscript, the authors used ChatGPT (OpenAI) for language editing and wording refinement to improve clarity and readability. The tool was not used for study design, data collection, data extraction, statistical analysis, interpretation of results, generation of references, or clinical conclusions. No identifiable patient-level data were entered into the tool. All AI-assisted edits were reviewed, verified, and approved by the authors, who take full responsibility for the integrity and content of the manuscript.

## Supplemental Material

Table S1: Electronic medical record data sources used for stroke eFI derivation

Table S2: Definitions of candidate deficit variables for electronic frailty index

Table S3: Full Multivariable Piecewise Cox Proportional Hazards Model for All-Cause Post-discharge Mortality

Table S4: Intravenous thrombolysis sub-cohort treatment outcomes stratified by frailty status

Figure S1: Calculation of the electronic frailty index

Figure S2: Relationship between age and electronic frailty index by sex

Figure S3: Frequency distribution of electronic frailty index scores

## Non-standard Abbreviations and Acronyms

ADL: activities of daily living
AI: artificial intelligence
AIS: acute ischemic stroke
aOR: adjusted odds ratio
BMI: body mass index
CFS: Clinical Frailty Scale
CKD: chronic kidney disease
ECASS III: European Cooperative Acute Stroke Study III
eFI: electronic frailty index
EMR: electronic medical record
EVT: endovascular thrombectomy
FI: frailty index
GI: gastrointestinal
GVIF: generalized variance inflation factor
HPB: hepatobiliary
ICD-10: International Classification of Diseases, 10th Revision
LACI: lacunar infarct
LOS: length of stay
mRS: modified Rankin Scale
MSW: medical social worker
mTICI: modified Treatment in Cerebral Infarction
NIHSS: National Institutes of Health Stroke Scale
OCSP: Oxfordshire Community Stroke Project
PACI: partial anterior circulation infarct
PH: parenchymal hematoma
POCI: posterior circulation infarct
RRR: relative risk ratio
TACI: total anterior circulation infarct
TOAST: Trial of Org 10172 in Acute Stroke Treatment

